# Evaluation of Simple and Convenient Methods for SARS-CoV-2 Detection in Wastewater in high and Low Resource Settings

**DOI:** 10.1101/2022.12.31.22284093

**Authors:** Pengbo Liu, Lizheng Guo, Matthew Cavallo, Caleb Cantrell, Stephen Patrick Hilton, Jillian Dunbar, Robbie Barbero, Robert Barclay, Orlando III Sablon, Marlene Wolfe, Ben Lepene, Christine Moe

## Abstract

Severe acute respiratory syndrome coronavirus-2 (SARS-CoV-2) RNA monitoring in wastewater has become an important tool for COVID-19 surveillance. Although many viral concentration methods such as membrane filtration and skim milk are reported, these methods generally require large volumes of wastewater, expensive lab equipment, and laborious processes. We utilized a Nanotrap^®^ Microbiome A Particles (Nanotrap particle) method for virus concentration in wastewater. The method was evaluated across six parameters: pH, temperature, incubation time, wastewater volumes, RNA extraction methods, and two virus concentration approaches vs. a one-step method. The method was further evaluated with the addition of the Nanotrap Enhancement Reagent 1 (ER1) by comparing the automated vs. a manual Nanotrap particle method. RT-qPCR targeting the nucleocapsid protein was used for detection and quantification of SARS-CoV-2 RNA. Different pH, temperature, incubation time, wastewater volumes, and RNA extraction methods did not result in reduced SARS-CoV-2 detection in wastewater samples. The two-step concentration method showed significantly better results (*P*<0.01) than the one-step method. Adding ER1 to wastewater prior to viral concentration using the Nanotrap particles significantly improved PCR Ct results (*P*<0.0001) in 10 mL grab samples processed by automated Nanotrap particle method or 10 mL and 40 mL samples processed by manual Nanotrap particle method. SARS-CoV-2 detection in 10 mL grab samples with ER1 and the automated method showed significantly better (*P*=0.0008) results than 150 mL grab samples using the membrane filtration method. SARS-CoV-2 detection in 10 mL swab samples with ER1 via the automated method was also significantly better than without ER1 (*P*<0.0001) and the skim milk method in 250 mL Moore swab samples (*P*=0.012). These results suggest that Nanotrap methods could substitute the traditional membrane filtration and skim milk methods for viral concentration without compromising on the assay sensitivity. The manual method can be used in resource-limited areas, and the high-throughput platform is appropriate for large-scale COVID-19 wastewater-based surveillance.

## INTRODUCTION

Coronavirus disease 2019 (COVID-19) is caused by severe acute respiratory syndrome coronavirus-2 (SARS-CoV-2), a single-stranded RNA virus that can infect individuals who can develop illness ranging in severity from life-threatening complications to mild symptomatic or asymptomatic infections. SARS-CoV-2 is mainly transmitted among people via respiratory droplets. However, it is also shed in feces at high concentrations and SARS-CoV-2 RNA titer in feces has been reported to be 10^5^ copies per gram of feces or between 10^2^-10^7^ genome copies per milliliter of stool suspension ^[1]^, allowing the virus RNA to be detected in sewage samples collected from wastewater treatment plants, community manholes, or buildings ^[2, 3]^. Since the early reported detection of SARS-CoV-2 in wastewater ^[4, 5]^, monitoring for SARS-CoV-2 RNA has become a critical tool for global COVID-19 surveillance and to guide response to COVID-19 outbreaks in communities. The initial step in processing wastewater samples for SARS-CoV-2 detection is to concentrate viruses from a relatively large volume to a small volume or pellet that can be used for nucleic acid extraction. Previous studies reported that the typical volumes of wastewater samples for the detection of SARS-CoV-2 range from 50-250 mL ^[6]^. For this range of volumes, SARS-CoV-2 was concentrated using several approaches: membrane filtration ^[4]^, precipitation with polyethylene glycol (PEG) ^[7]^, skim milk ^[8]^, ultracentrifugation ^[9]^, and ultrafiltration, ^[10]^etc. This step usually requires a centrifuge that can spin down at least a 50-mL tube, which is a problem for some laboratories with limited resources in the world. Using a small volume of wastewater samples without this centrifuge and without compromise on assay sensitivity would be preferable for broad application of SARS-COV-2 wastewater surveillance. In addition, there are limited reports for application of novel viral concentration approach in wastewater samples and consideration of small sample volume with better sensitivity. Nanotrap particles are highly porous hydrogel particles that are versatile in their functionality for the capture and concentration of analytes, such as proteins, peptides, nucleic acids, hormones, viral antigens, and live infectious pathogens. This versatility allows Nanotrap particles to be customized to capture analytes in various complex biological mixtures like blood, saliva, urine, and wastewater ^[11]^. Nanotrap particles can perform three essential functions: capturing target analytes from complex metrics, separating analytes from interfering materials, and protecting target analytes from degradation. Nanotrap particles are capable of preserving viruses and nucleic acids from degradation after sample concentration at both ambient and increased temperature for up to 3 days ^[12]^. In addition to these advantages, Nanotrap particles can capture and concentrate multiple pathogens in one sample ^[13]^. This capability has been demonstrated with Nanotrap particles used in a coinfection scenario with HIV and Rift Valley Fever Virus in bovine serum ^[12]^. Since the COVID-19 pandemic began, Nanotrap particles have been used to detect SARS-CoV-2 ^[14, 15]^ and other respiratory viruses ^[16, 17]^ in wastewater. For example, Anderson et al ^[17]^ demonstrated that Nanotrap particles can be used for simultaneous concentration and detection of SARS-CoV-2, influenza, and Respiratory Syncytial Virus in wastewater.

Thee sampling methods were used for SARS-CoV-2 wastewater-based epidemiology: grab, composite, and passive sampling. Grab sampling is a simple and convenient method that wastewater is collected at one point in time; however, interpretation of the results is limited because the samples only represent a snapshot at one moment. Composite sampling has been considered as a more representative method due to its ability to collect numerous individual samples at regular time intervals, and the individual samples are subsequently combined in proportion to the wastewater flow rate. However, composite sampling is more costly and time-consuming, and it may not be feasible under certain environmental conditions. Passive sampling, which a material is deployed in wastewater to trap SARS-CoV-2 over time, provides a sensitive, low-cost, and convenient alternative to composite sampling. We used a Moore swab passive sampling method, pieces of gauzes tied to a fishing line for SASR-CoV-2 wastewater surveillance in institutional buildings. Grab samples were collected from manholes of the buildings and membrane filtration method was used to concentrate the virus from 80-500 mL wastewater because the grab samples usually had lower turbidity. Moore swab samples were placed in the stream of wastewater and collected after 24 hours. Because these samples had higher turbidity, the virus was concentrated using the skim milk method.

In this study, we evaluated whether some common parameters, such as pH, temperature, incubation time, wastewater volume, RNA extraction method, and two-step (skim milk + Nanotrap particle method) vs. one-step (Nanotrap particle method alone) concentration methods impact the detection of SARS-CoV-2 RNA in wastewater. The secondary objectives were to compare: a) optimized high-throughput, automated sample processing vs. manual Nanotrap particle methods with a small sample volume (10 mL) so we can scale up wastewater surveillance if automated sample processing shows at least similar sensitivity as the manual method, and b) compare manual Nanotrap concentration methods (for 10 mL) to membrane filtration method for grab samples (150 mL) and skim milk method for Moore swab samples (250 mL) so we can use this method in settings with low resources. We would like to ensure that switching concentration methods and sample volumes from traditional membrane filtration and skim milk methods to Nanotrap method do not compromise the sensitivity.

## MATERIALS and METHODS

### Wastewater samples

For this study, wastewater samples were collected weekly from community manholes and influent lines of wastewater treatment plants in Atlanta from June 2021 to August 2021. Samples from community manholes were collected weekly using Moore swabs made from cotton gauze and secured by typing the fishing line to a hook at the top of the manhole and the swabs were placed in wastewater flows for 24 hours and then retrieved.

Samples from influent lines were 500 mL grab samples collected in wastewater treatment plants in urban Atlanta areas, which represented tens of thousands of people. Sample collection procedures are described in detail at https://www.protocols.io/view/wastewater-sample-collection-moore-swab-and-grab-s-b2rzqd76

### Membrane filtration

The wastewater samples were centrifuged at 5,000 rpm for 5 minutes at 4°C to remove solids in the sample that could clog the membrane filter. The method using 0.45-*µ*m-pore-size, 47-mm-diameter nitrocellulose filters, described by Liu et al^[3]^, was used to concentrate SARS-CoV-2 from 150 mL of wastewater grab samples after pH adjustment to 3.5 and 25 mM of magnesium chloride was added. Before filtration, 10^5^ equivalent genome copies (EGC) of Bovine Respiratory Syncytial Virus (BRSV) (INFORCE 3, Zoetis, Parsippany, NJ) were added to the sample as a process control. After filtration, the membrane filter was placed into a microcentrifuge tube and 800 *µ*L of RLT buffer from the RNeasy Mini Kit (QIAGEN, Hilden, Germany) was added immediately. The sample was vortexed at maximum speed for 10 minutes and then subjected to RNA extraction as described in the instructions of the RNeasy Mini Kit.

### Skim milk flocculation concentration

Skim milk flocculation method, described by Liu et al ^[3]^ was used to concentrate SARS-CoV-2 from Moore swab samples collected from community manholes in Atlanta. Briefly, a 5% (w/v) skimmed milk solution was prepared by dissolving 5 g of skimmed milk powder (BD, #232100, Sparks, MD) in 100 mL of distilled water. Before the flocculation step, the pH of the liquid sample squeezed from Moore swab was adjusted to 3.5 using 6 N HCL and then skimmed milk was added at a final concentration of 1%, followed by addition of 10^5^ EGC of BRSV and shaking for 2 h (https://www.protocols.io/view/skim-milk-flocculation-and-rna-extraction-for-sars-b2uwqexe).

### Manual Nanotrap particle method for concentration of SARS-CoV-2 and RNA extraction

Nanotrap Microbiome A Particles (SKU#44202, Ceres Nanosciences Inc., Manassas, VA) are magnetically functionalized, affinity capture hydrogel particles that capture and concentrate microbes from samples. Grab wastewater samples from influent lines or of the liquid squeezed from Moore swab samples from community manholes were used for the manual Nanotrap particle method. Four-hundred microliters of Nanotrap^®^ particles and ten microliters of BRSV, approximately equivalent to 10^5^ EGC of BRSV, were added to 10 mL or 40 mL of wastewater. Seeded samples were incubated for 20 minutes and were placed on a magnet rack (Thermo Fisher Scientific, Waltham, MA) for 10 minutes. Supernatant was then removed without disturbing the pellet of Nanotrap^®^ particles. One milliliter of molecular grade water was used to rinse the pellet off the side of the tube and the sample was transferred to a 1.7 mL tube. The sample was then placed on a small magnet rack for 2 minutes to allow the particles to pellet, followed by adding 140 *µ*L of 1×PBS to the particle pellet that was used for RNA extraction using the QIAamp Viral RNA Mini Kit (QIAGEN, Valencia, CA) in accordance with the manufacturer’s instructions. (https://www.protocols.io/view/manual-nanotrap-concentration-and-rna-extraction-f-b2uzqex6).

### Automated Nanotrap particle method for SARS-CoV-2 concentration and RNA extraction

KingFisher^™^ Apex robot platform (Thermo Fisher Scientific, USA), allowing to process 24 samples using a 24-plex head, was used for virus concentration and nucleic acid extraction from wastewater. Briefly, 10 *µ*L of BRSV (equivalent to 10^5^ EGC of BRSV) and 50 *µ*L of Nanotrap^®^ enhancement reagent 1 (ER1, Ceres Nanoscience Inc., #10111) were mixed with approximate 5 mL of wastewater and two replicate wells with a total of 10 mL of wastewater were used for each sample. After 10 min of incubation at room temperature, 75 *µ*L of Nanotrap^®^ particles were added and the sample plates were loaded into the KingFisher^™^ Apex, followed by running the designated KingFisher^™^ script. After viral concentration, samples were processed for RNA extraction using the Applied Biosystems MagMax™ nucleic acid isolation kit (Thermo Fisher Scientific #48310) on the same platform following the instructions. (https://www.protocols.io/view/nanotrap-kingfisher-concentration-extraction-amp-m-b2nkqdcw).

### Quantitative real-time RT-PCR method

SARS-CoV-2 RNA was detected via RT-qPCR (https://www.protocols.io/view/singleplex-qpcr-for-sars-cov-2-n1-and-brsv-b2qyqdxw) using the N1 primers described before ^[18, 3]^ and the TaqPath™ qPCR Master Mix (ThermoFisher Scientific, Waltham, MA). Synthetic SARS-CoV-2 RNA (ATCC VR-3276SD, Manassas, VA) with known concentration was 10-fold diluted and the standard curve was used for quantification of SARS-CoV-2 RNA titers in wastewater. Potential PCR inhibition was examined in some samples through testing dilutions (1:2, 1:5 and 1:10) and comparing the results to those from the undiluted RNA. Positive samples were defined as the presence of Ct values in both duplicate wells from one sample; if both Ct values were below 36 with the Ct difference <2, we considered the samples as positive; if both Ct values were above 36, the sample was classified as weak positive; if both or either Ct were absent or greater than 36, the sample was classified as negative.

### Statistical analysis

A paired nonparametric Friedman test was used to compare the mean Ct values of the groups of Nanotrap particle characterization experiments. Wilcoxon signed rank test was used to compare the two-step vs. one-step concentration methods, the automated vs. manual Nanotrap particle methods with and without ER1 in 10 mL and 40 mL grab and Moore swab samples, the automated Nanotrap particle method vs. the membrane filtration method in grab samples, and the automated Nanotrap particle method vs. the skim milk method in Moore swab samples. Differences were considered statistically significant if the p-value was <0.05.

## RESULTS

We first evaluated the effects of four physical and chemical parameters (incubation time, temperature, pH, and sample volume) on the manual Nanotrap particle method without ER1 (Figure 1a-1d). Incubation of 10 mL wastewater with 140 *µ*L Nanotrap particles at different temperatures (4°C, 22°C, 37°C, and 50°C) for 20 mins followed by RNA extraction and detection of SARS-CoV-2 RNA using RT-qPCR did not show a difference (*P*>0.05) in Ct values (Figure 1a). Incubation of 10 mL wastewater samples with Nanotrap particles at room temperature for different times (10, 20, 40, 60 minutes) did not show a significant difference (*P*>0.05) in Ct values for SARS-CoV-2 detection (Figure 1b). Wastewater adjusted to pH values of 5.0, 8.0, and 10.0 at room temperature before sample processing was not associated with differences in SARS-CoV-2 Ct values in 10 mL wastewater samples using Nanotrap particles (Figure 1c). Ct values of SARS-CoV-2 detection in 10 mL, 40 mL and 120 mL wastewater samples showed no statistical difference (*P*>0.05) (Figure 1d). These results suggest that Nanotrap particles are very stable and are environmentally tolerant, which is appropriate for application in other countries and is convenient for transportation and storage.

**Figure 1.**
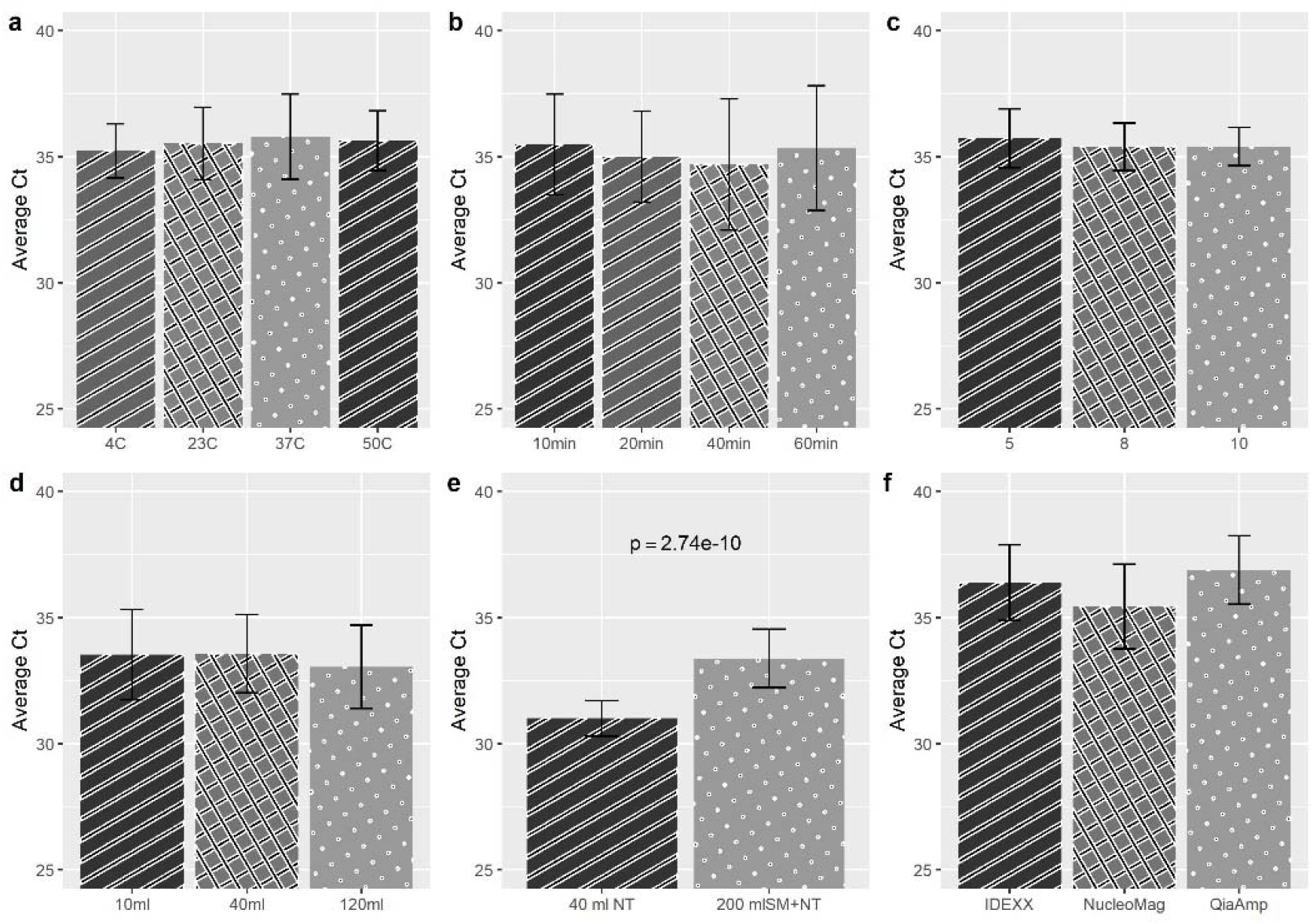
Nanotrap Microbiome A Particles captured SARS-CoV-2 from wastewater. a) Incubation temperature (4°C, 23°C, 37°C, and 50°C) of wastewater samples with Nanotrap particles; b) Incubation time (10 min, 20 min, 40 min, and 60 min) of wastewater samples with Nanotrap particles; c) pH (5, 8, and 10) of wastewater sample; d) Processing sample volumes (10 mL, 40 mL, and 120 mL) with SARS-CoV-2 detection; e) Two-step concentration method (skim milk + Nanotrap particle method) for 200 mL wastewater samples compared to Nanotrap particle method alone for 40 mL samples showed significance difference when compared two-step vs. one-step using Wilcoxon signed rank test; f) Comparison of three RNA extraction methods (IDEXX, MagMax and QIAamp) *Four samples tested in duplicate experiments with duplicate wells in RT-qPCR yielded 16 data points for each parameter evaluation

We then compared a two-step concentration method that started with skim milk concentration followed by Nanotrap particle concentration for 200 mL wastewater samples to the one-step Nanotrap particle method for 40 mL wastewater samples. We observed significantly lower SARS-CoV-2 Ct values (*P*<0.0001) with the two-step method and a larger sample volume (Figure 1e), suggesting that two-step of viral concentration with large volume is more sensitive for viral detection than one step concentration from a small volume of wastewater. Finally, we compared three RNA extraction kits (QIAamp, MagMAX and IDEXX kits) to extract SARS-CoV-2 RNA from 10 mL same wastewater samples for each kit followed by RT-qPCR detection and observed that these gave similar results (*P*=0.103) (Figure 1f). These results indicate that the three RNA extraction methods are same sensitive for viral detection.

The effect of ER1 for the Nanotrap particles was examined by comparing the automated Nanotrap particle method with and without ER1 and the manual Nanotrap particle method with two sample volumes (10 mL and 40 mL) with and without ER1 using wastewater grab samples from influent lines (Figure 2).

**Figure 2.**
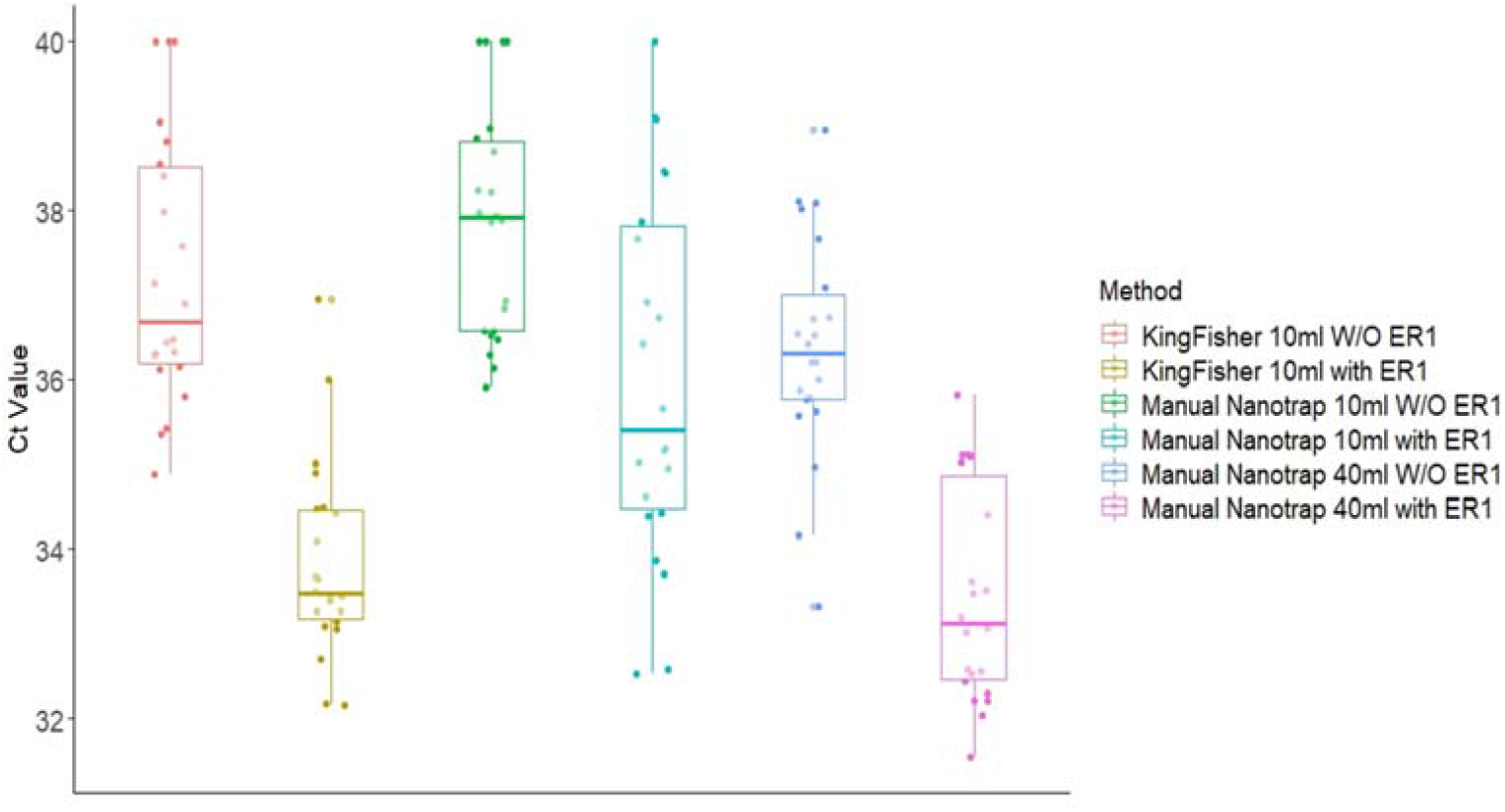
Comparison of SARS-CoV-2 RNA detection in 10 mL grab wastewater samples from influent lines processed using the automated Nanotrap particle method and the manual Nanotrap particle method (10 mL and 40 mL) with and without ER1.

Adding ER1 to 10 mL grab wastewater samples prior to viral concentration on the KingFisher system resulted in significantly lower (*P*=0.0005) Ct values compared to the protocol without ER1. When the manual Nanotrap particle method was used, adding ER1 to both 10 mL and 40 mL grab samples also showed a similar improvement (*P*>0.05) in Ct values (Figure 2). These results suggest that including ER1 in the viral concentration significantly increase the viral detection in in grab samples using both the manual and the automated Nanotrap assays.

We then examined the effect of ER1 using Moore swab samples and again compared the automated Nanotrap particle method (10 mL sample) and manual Nanotrap particle method (10 mL and 40 mL) with and without ER1 (Figure 3). Adding ER1 to the liquid squeezed from 10 mL Moore swab samples prior to viral concentration with Nanotrap particles yielded significantly lower Ct values (*P*<0.05) in the automated Nanotrap particle method. Adding ER1 to 10 mL and 40 mL Moore swab samples and using the manual Nanotrap particle method also demonstrated lower Ct values compared to samples without ER1, but the Ct values from the Moore swab samples were more variable than those from the grab samples (Figure 3). These results indicate that ER1 significantly increase viral concentration in Moore swab samples using both the manual and the automated Nanotrap assays

**Figure 3.**
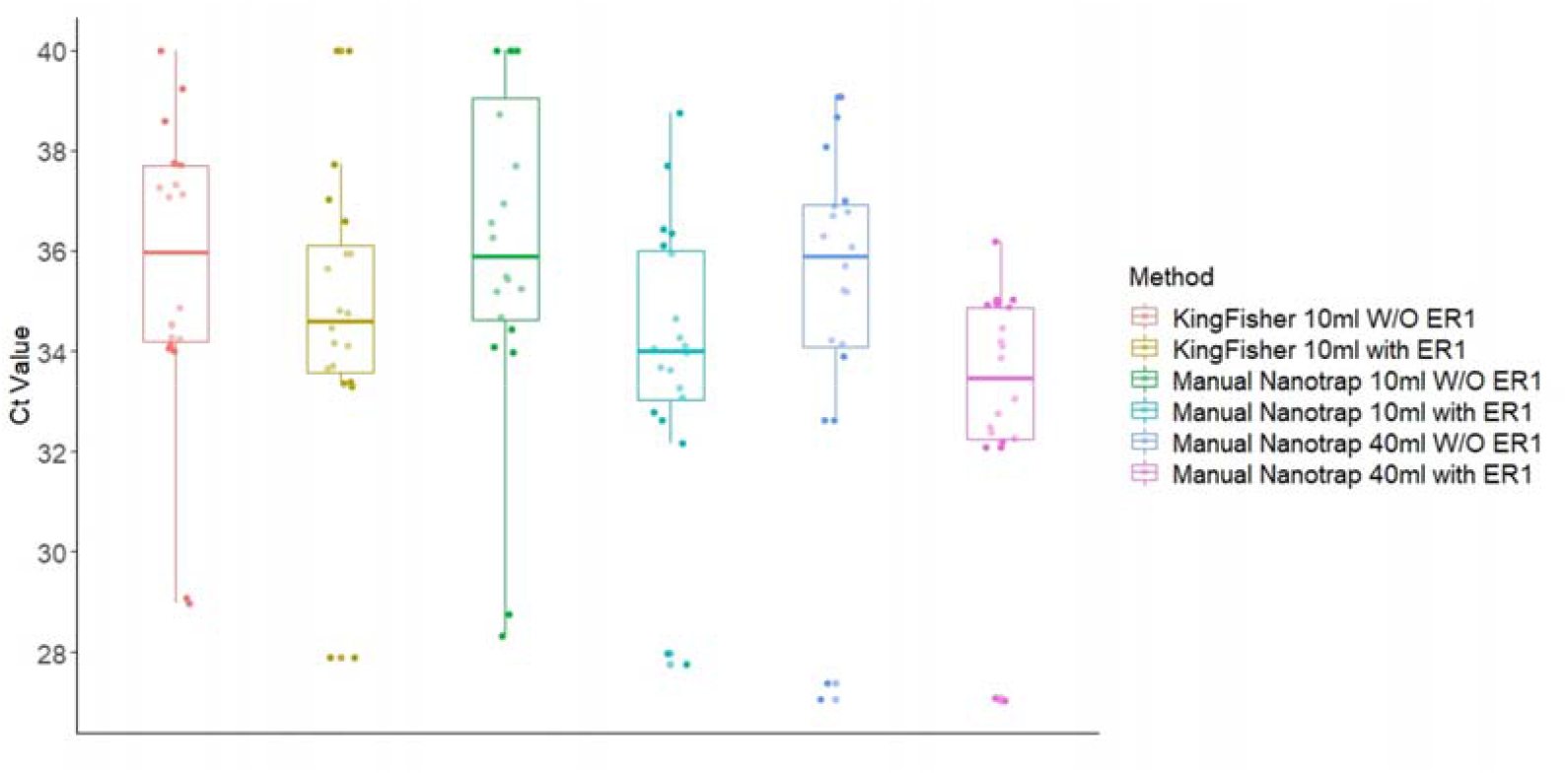
Comparison of SARS-CoV-2 RNA detection in Moore swab samples processed using the automated Nanotrap particle method (10 mL) and the manual Nanotrap particle method (10 mL and 40 mL) with and without adding ER1.

Typically, virus detection in environmental samples requires concentration from large sample volumes which can be more challenging to process with a high-throughput platform. We compared the automated Nanotrap particle method and ER1 using 10 mL wastewater samples to two traditional concentration methods that use larger volume samples: the membrane filtration method using 150 mL grab samples (Figure 4 and Table 1) and the skim milk method using 250 mL Moore swab samples (Figure 5 and Table 2).

**Figure 4.**
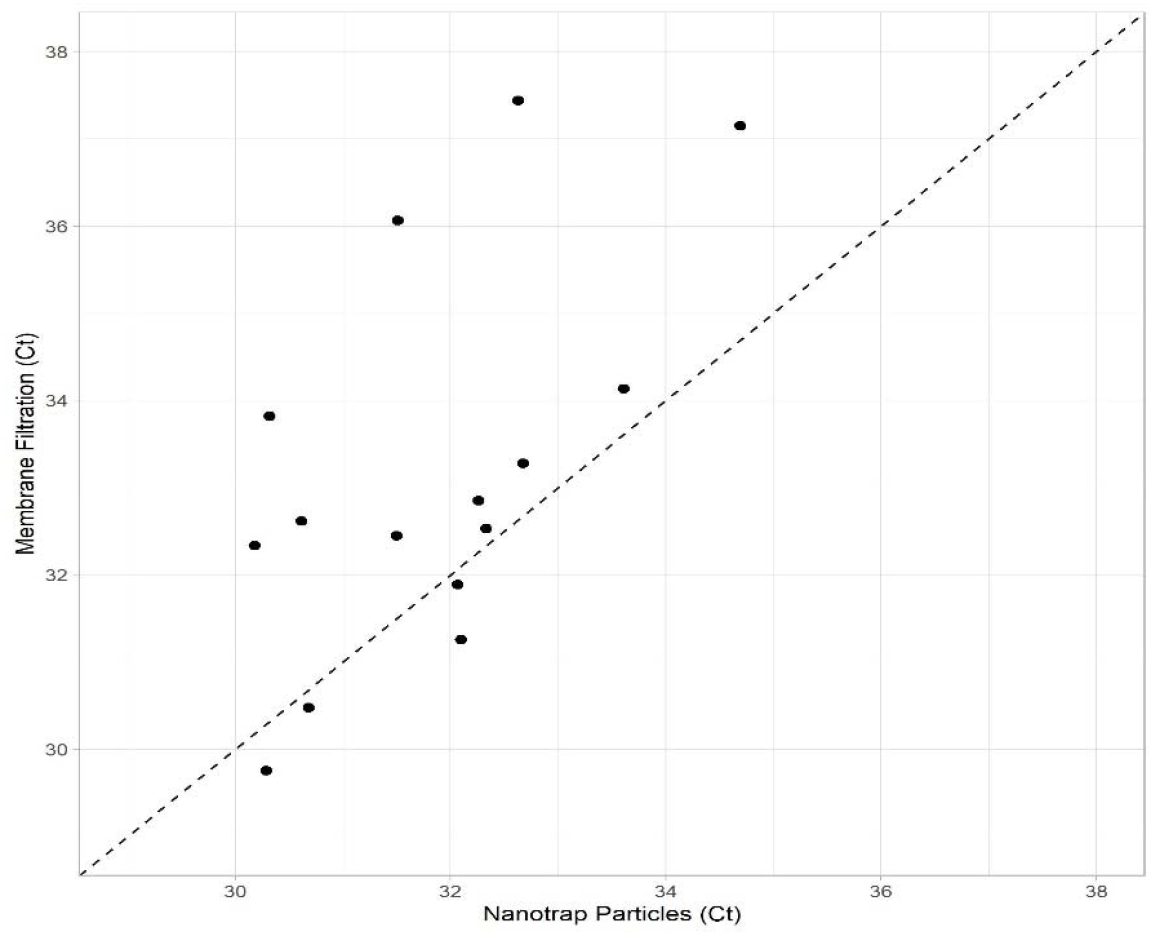
Comparison of automated Nanotrap particle method and ER1 for processing 10 mL grab wastewater with vs. membrane filtration method for processing of 150 mL wastewater in detection of SARS-CoV-2 RNA.

**Table 1.** Comparison of SARS-CoV-2 detection in 10 mL grab wastewater using the automated Nanotrap particle method with and without ER1 vs. membrane filtration method in 150 mL wastewater.

| Method | Sample Number | Positive (%) | Average Ct (SD*) |
| --- | --- | --- | --- |
| Automated Nanotrap particle method w/ ER1 | 15 | 15 (100) | 31.55 (1.15) |
| Automated Nanotrap particle method w/o ER1 | 15 | 15 (100) | 35.60 (1.13) |
| Membrane Filtration | 15 | 15 (100) | 32.78 (1.96) |

\*Standard deviation
Wilcoxon Signed Rank Test:
Automated Nanotrap particle method with ER1 vs. Automated Nanotrap particle method w/o ER1, $P < 0.0001$
Automated Nanotrap particle method with ER1 vs. membrane filtration, $P = 0.0083$
Automated Nanotrap particle method w/o ER1 vs. membrane filtration, $P = 0.0008$

**Figure 5.**
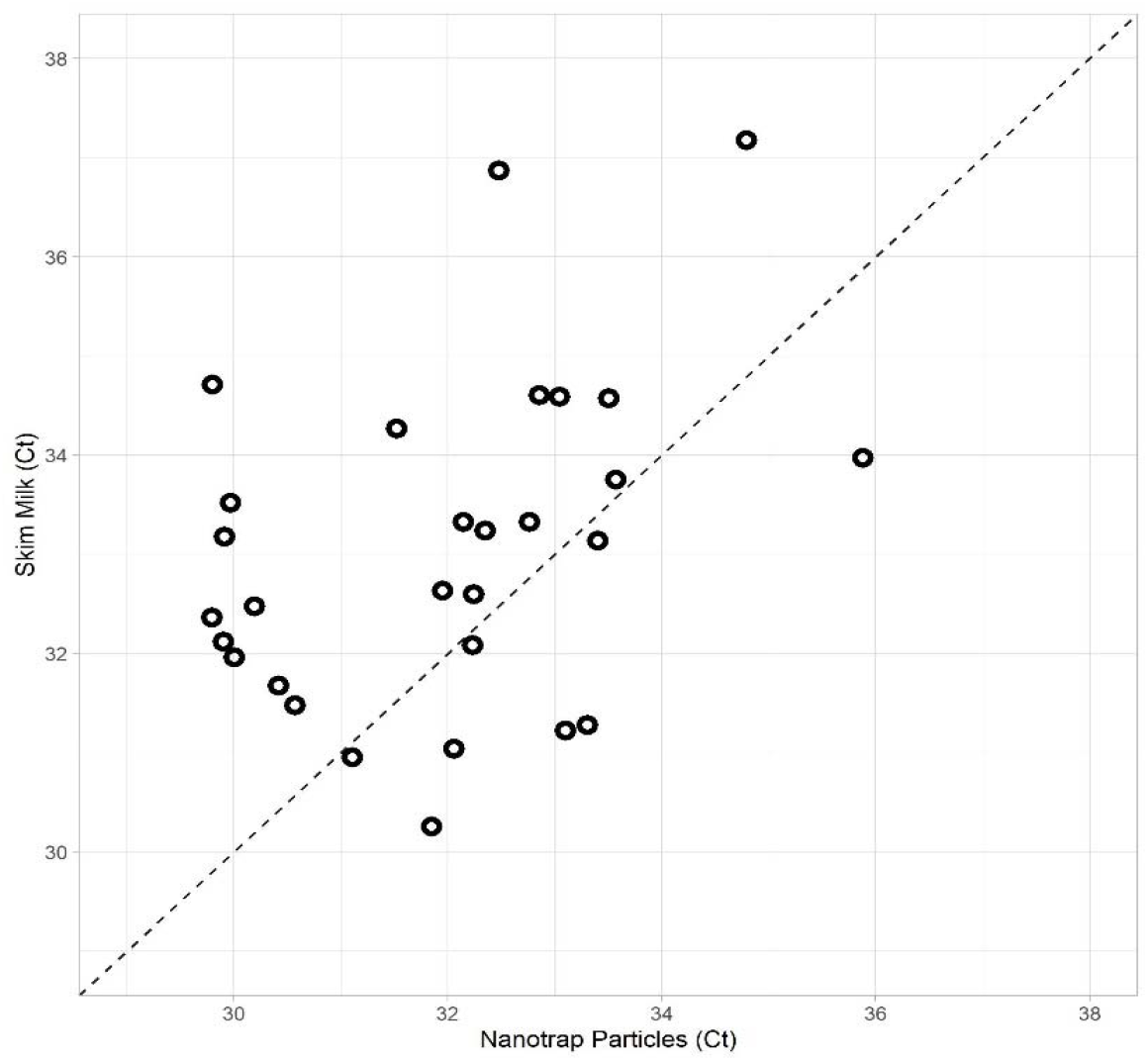
Comparison of automated Nanotrap particle method for processing 10 mL Moore swab wastewater vs. skim milk method for processing of 250 mL Moore swab wastewater for detection of SARS-CoV-2 RNA.

**Table 2.** Comparison of SARS-CoV-2 RNA detection in 10 mL Moore swab samples using the automated Nanotrap particle method and membrane filtration method for processing of 150 mL

| Method | Sample Number | Positive (%) | Average Ct (SD*) |
| --- | --- | --- | --- |
| Automated Nanotrap particle method with ER1 | 29 | 29 (100) | 31.92 (1.60) |
| Automated Nanotrap particle method w/o ER1 | 29 | 29 (100) | 34.17 (2.01) |
| Skim milk | 29 | 29 (100) | 32.97 (1.58) |

\*Standard deviation
Wilcoxon Signed Rank Test:
Automated Nanotrap particle method with ER1 vs. w/o ER1, $P < 0.0001$
Automated Nanotrap particle method with ER1 vs. skim milk, $P = 0.006$
Automated Nanotrap particle method w/o ER1 vs. skim milk, $P = 0.012$

Fifteen wastewater grab samples were processed in parallel using the automated Nanotrap particle method with ER1 (10 mL) and without ER1 (10 mL), as well as by the membrane filtration method (150 mL). In comparison to without ER1, adding ER1 to Nanotrap particles in wastewater significantly improved SARS-CoV-2 RNA detection in 10 mL wastewater by an average of 4.05 Ct values (*P*<0.0001). Although all 15 samples were positive by both methods, SARS-CoV-2 detection in 10 mL grab samples with ER1 and Nanotrap particles showed significantly lower average Ct values (*P*=0.0083) than the membrane filtration method using 150 mL grab sample. SARS-CoV-2 detection in 10 mL sample without ER1 was also significantly better (*P*=0.0008) than the membrane filtration method using 150 mL grab sample (Table 1). These results suggest that using Nanotrap particles to concentrate viruses from small volume of grab samples can achieve even better results than using membrane filtration method in large volume wastewater.

Similarly, wastewater extracted from 29 Moore swab samples was processed side-by-side using the automated Nanotrap particle method with ER1 with 10 mL samples and without ER1 in 10 mL samples vs. using the skim milk method with 250 mL samples. SARS-CoV-2 RNA detection in 10 mL swab sample with the automated Nanotrap particle method and ER1 was statistically significantly better (*p*<0.0001) than the automated Nanotrap particle method without ER1 when Ct values were compared. In addition, the average Ct value for the 10 mL samples processed using the Nanotrap particles were significantly lower (*p*=0.006) than average Ct values for the skim milk method using 250 mL samples (Table 2 and Figure 5). These results indicate that using Nanotrap particles to concentrate viruses from small volume of Moore swab samples can achieve significant better results than using skim milk method in large volume swab samples.

## DISCUSSION

Since the COVID-19 pandemic started, many studies have focused on the detection of SARS-CoV-2 RNA in wastewater collected from wastewater treatment plants, community manholes, campus residence halls and other buildings. Although infected human subjects shed a relatively high titer of SARS-CoV-2 in their feces, the viruses can become highly diluted in wastewater and are generally present in low concentration in wastewater ^[19]^, especially when case numbers are lower in communities. To detect SARS-CoV-2 RNA in wastewater, investigators tend to concentrate the viruses from large volumes ^[20, 10, 21]^ into smaller volumes or precipitate the viruses into a pellet ^[22]^ so that nucleic acid amplification and detection methods such as RT-qPCR can be applied. For this reason, most studies describe one or two in-series methods for viral concentration in wastewater samples. These methods include adsorption and elution ^[20]^, polyethylene glycol (PEG) ^[7]^, skim milk ^[8]^, membrane filtration ^[4]^, ultracentrifugation ^[9]^, ultrafiltration^[10]^, bag-mediated filtration system ^[21]^,etc. When considering these concentration methods, factors such as sample volume, turbidity, and required lab equipment are critical since these factors affect the efficiency of virus concentration, RNA extraction, and subsequent RT-qPCR detection, as well as the feasibility of analyzing large numbers of samples on a routine basis. Another important consideration with molecular detection of viral RNA in wastewater samples is that certain organic matter and chemicals that can inhibit RT-PCR reactions can be concentrated along with SARS-CoV-2 and negatively affect the RT-qPCR results. These inhibitors can cause a weaker PCR signal or even false negative results. Due to these limitations of the aforementioned concentration methods, there is a need for a simple, rapid, robust, and efficient concentration method that can be automated for large-scale COVID-19 wastewater surveillance or that can be performed manually in resource limited areas.

In this study, we first evaluated the robustness of the Nanotrap particle methods across six parameters (pH, temperature, incubation time, wastewater volumes, RNA extraction methods, and two virus concentration approaches: a two-step process vs. a one-step method). Different pH, temperature, incubation time, wastewater volumes, and RNA extraction methods did not result in reduced SARS-CoV-2 detection in wastewater samples, demonstrating the robustness of the Nanotrap particle methods for virus capture from wastewater. Adding a skim milk flocculation step from a larger sample volume prior to the Nanotrap particle method offered significantly better results than the Nanotrap particle method by itself. These results indicate that virus concentration methods using Nanotrap particles from large volumes might be beneficial during times when lower number of COVID-19 cases are reported and greater detection sensitivity is required.

We utilized a Nanotrap particle method for virus concentration in 10 mL wastewater samples using: 1) a KingFisher Apex system for automated virus concentration and RNA extraction, and 2) a manual viral concentration method using a magnetic tube rack. SARS-CoV-2 RNA was extracted using an Applied Biosystems MagMax nucleic acid isolation kit. Subsequently, RT-qPCR with primers and probes targeting the nucleocapsid protein was used for detection and quantification of SARS-CoV-2 RNA. Our results indicated that adding ER1 to wastewater prior to viral concentration using the Nanotrap particles significantly improved PCR results in 10 mL samples processed in an automated method or in 10 ml and 40 mL samples processed using the manual method compared to not using ER1 for the same wastewater samples. We noted generally increasing Ct values by 3.05 for grab samples and 2.25 for swab samples compared to the same sample and the same method without ER1. In addition, SARS-CoV-2 RNA detection in 10 mL grab samples with Nanotrap particles and ER1 showed significantly better results than 150 mL grab samples using the membrane filtration method and 250 mL Moore swab samples using the skim milk method.

Compared to traditional pathogen concentration methods such as skim milk and membrane filtration methods (Table 3), concentrating viruses from wastewater using Nanotrap particles has several advantages: 1) small sample volume (10 mL), which is easier to collect and transport; 2) simple equipment, only requiring a magnetic tube rack, which is appropriate for low-resource settings; 3) potential to adapt to high throughput platform for scalable implementation; 4) more sensitive than traditional large volume concentration methods (membrane filtration and skim milk methods); and 5) rapid – viral concentration takes significantly less than an hour and requires no additional centrifugation or filtrations steps for both the high throughput and manual methods; 6) long shelf life of Nanotrap particles which makes easy for storage and transportation. All of these advantages enable this method to be used in resource-limited areas and allow SARS-CoV-2 wastewater surveillance to be implemented via an efficient and scalable approach.

**Table 3.** Comparison of Nanotrap Microbiome A Particles, skim milk, and membrane filtration methods for SARS-CoV-2 detection in wastewater

|  | Nanotrap Microbiome A Particles |  | Skim Milk | Membrane Filtration |
| --- | --- | --- | --- | --- |
|  | KingFisher Apex | Manual |  |  |
| **Cost (\$)/per Sample\*** | 14.8 | 12.0 | 8.3 | 10.0 |
| **Processing Time (hrs) for 10 samples\*** | 2.5 | 3.0-3.5 | 4.0-5.0 | 4.0-6.0 |
| **Sample Volume (mL)** | 10 | 10 or 40 | 250 | 150 |
| **Sample Type** | Grab or Swab | Grab or swab | Better for swab samples | Better for grab samples |
| **Advantage** | - Automatic<br>- Less labor<br>- Small sample volume<br>- Sensitive<br>- Can be used on turbid and clear water samples<br>- Only a microcentrifuge required | - Easy to use and not a long protocol<br>- Stable to store and transport<br>- Small volume of sample for high sensitivity<br>- Can be used on turbid and clear water samples<br>- Only a microcentrifuge required<br>- Affordable | - Affordable<br>- Low number of consumables used | - Affordable<br>- Only a microcentrifuge required<br>- Low number of consumables used |
| **Disadvantage** | - Expensive equipment<br>- More plastic consumable |  | - Requires centrifuge if large volume is used<br>- More time consuming for flocculation<br>- Not sensitive compared with the Nanotrap method | - Filtration may take more time if water sample is turbid<br>- Not sensitive compared with the Nanotrap method |

\*Including both virus concentration and RNA extraction

## Data Availability

All data produced in the present study are available upon reasonable request to the authors

## Acknowledgements

We thank the financial support from the NIH Rapid Acceleration of Diagnostics (RADx) initiative (contract No. 75N92021C00012 to Ceres Nanosciences, Inc). We are debt to Salvins J. Strods, Chipo Chemoyo J. Baker Afamefuna from RADx Initiative and Dr. Louis J. Vuga from NIH. We appreciate all of the wastewater collection support from Jamie VanTassell (Emory University), Lorenzo Freeman, Wayne Rose and Carl Holt at the City of Atlanta Department of Watershed Management.

## References

1. Jones DL, Baluja MQ, Graham DW, Corbishley A, McDonald JE, Malham SK, et al. Shedding of SARS-CoV-2 in feces and urine and its potential role in person-to-person transmission and the environment-based spread of COVID-19. Sci Total Environ. 2020 Dec 20;749:141364.

2. Gibas C, Lambirth K, Mittal N, Juel MAI, Barua VB, Roppolo Brazell L, et al. Implementing building-level SARS-CoV-2 wastewater surveillance on a university campus. Sci Total Environ. 2021 Aug 15;782:146749.

3. Liu P, Ibaraki M, VanTassell J, Geith K, Cavallo M, Kann R, et al. A sensitive, simple, and low-cost method for COVID-19 wastewater surveillance at an institutional level. Sci Total Environ. 2022 Feb 10;807(Pt 3):151047.

4. Ahmed W, Angel N, Edson J, Bibby K, Bivins A, O’Brien JW, et al. First confirmed detection of SARS-CoV-2 in untreated wastewater in Australia: A proof of concept for the wastewater surveillance of COVID-19 in the community. Sci Total Environ. 2020 Aug 1;728:138764.

5. Wu F, Zhang J, Xiao A, Gu X, Lee WL, Armas F, et al. SARS-CoV-2 Titers in Wastewater Are Higher than Expected from Clinically Confirmed Cases. mSystems. 2020 Jul 21;5(4).

6. Khan K, Tighe SW, Badireddy AR. Factors influencing recovery of SARS-CoV-2 RNA in raw sewage and wastewater sludge using polyethylene glycol-based concentration method. J Biomol Tech. 2021 Sep;32(3):172–79.

7. Wu F, Xiao A, Zhang J, Moniz K, Endo N, Armas F, et al. Wastewater surveillance of SARS-CoV-2 across 40 U.S. states from February to June 2020. Water Res. 2021 Sep 1;202:117400.

8. Philo SE, Ong AQW, Keim EK, Swanstrom R, Kossik AL, Zhou NA, et al. Development and Validation of the Skimmed Milk Pellet Extraction Protocol for SARS-CoV-2 Wastewater Surveillance. Food Environ Virol. 2022 Feb 10.

9. Wurtzer S, Marechal V, Mouchel JM, Maday Y, Teyssou R, Richard E, et al. Evaluation of lockdown effect on SARS-CoV-2 dynamics through viral genome quantification in waste water, Greater Paris, France, 5 March to 23 April 2020. Euro Surveill. 2020 Dec;25(50).

10. Fores E, Bofill-Mas S, Itarte M, Martinez-Puchol S, Hundesa A, Calvo M, et al. Evaluation of two rapid ultrafiltration-based methods for SARS-CoV-2 concentration from wastewater. Sci Total Environ. 2021 May 10;768:144786.

11. Jaworski E, Saifuddin M, Sampey G, Shafagati N, Van Duyne R, Iordanskiy S, et al. The use of Nanotrap particles technology in capturing HIV-1 virions and viral proteins from infected cells. PLoS One. 2014;9(5):e96778.

12. Shafagati N, Narayanan A, Baer A, Fite K, Pinkham C, Bailey C, et al. The use of NanoTrap particles as a sample enrichment method to enhance the detection of Rift Valley Fever Virus. PLoS Negl Trop Dis. 2013;7(7):e2296.

13. Lin SC, Carey BD, Callahan V, Lee JH, Bracci N, Patnaik A, et al. Use of Nanotrap particles for the capture and enrichment of Zika, chikungunya and dengue viruses in urine. PLoS One. 2020;15(1):e0227058.

14. Karthikeyan S, Nguyen A, McDonald D, Zong Y, Ronquillo N, Ren J, et al. Rapid, Large-Scale Wastewater Surveillance and Automated Reporting System Enable Early Detection of Nearly 85% of COVID-19 Cases on a University Campus. mSystems. 2021 Aug 31;6(4):e0079321.

15. Karthikeyan S, Ronquillo N, Belda-Ferre P, Alvarado D, Javidi T, Longhurst CA, et al. High-Throughput Wastewater SARS-CoV-2 Detection Enables Forecasting of Community Infection Dynamics in San Diego County. mSystems. 2021 Mar 2;6(2).

16. Shafagati N, Fite K, Patanarut A, Baer A, Pinkham C, An S, et al. Enhanced detection of respiratory pathogens with nanotrap particles. Virulence. 2016 Oct 2;7(7):756–69.

17. Anderson P BS, Barclay RA, Smith N, Fernandes J, Besse K, Goldfarb D, Barbero R, Dunlap R, et al. Nanotrap particles improve nanopore sequencing of SARS-CoV-2 and other respiratory viruses. bioRxiv Preprint. 2021 December 9, 2021.

18. Lu X, Wang L, Sakthivel SK, Whitaker B, Murray J, Kamili S, et al. US CDC Real-Time Reverse Transcription PCR Panel for Detection of Severe Acute Respiratory Syndrome Coronavirus 2. Emerg Infect Dis. 2020 Aug;26(8).

19. Yang S, Dong Q, Li S, Cheng Z, Kang X, Ren D, et al. Persistence of SARS-CoV-2 RNA in wastewater after the end of the COVID-19 epidemics. J Hazard Mater. 2022 May 5;429:128358.

20. Barril PA, Pianciola LA, Mazzeo M, Ousset MJ, Jaureguiberry MV, Alessandrello M, et al. Evaluation of viral concentration methods for SARS-CoV-2 recovery from wastewaters. Sci Total Environ. 2021 Feb 20;756:144105.

21. Philo SE, Keim EK, Swanstrom R, Ong AQW, Burnor EA, Kossik AL, et al. A comparison of SARS-CoV-2 wastewater concentration methods for environmental surveillance. Sci Total Environ. 2021 Mar 15;760:144215.

22. Wolfe MK, Topol A, Knudson A, Simpson A, White B, Vugia DJ, et al. High-Frequency, High-Throughput Quantification of SARS-CoV-2 RNA in Wastewater Settled Solids at Eight Publicly Owned Treatment Works in Northern California Shows Strong Association with COVID-19 Incidence. mSystems. 2021 Oct 26;6(5):e0082921.

